# Implementation of Agile in health care: Methodology for a multi-site home hospital accelerator

**DOI:** 10.1101/2023.12.12.23299864

**Authors:** Meghna Desai, Miriam Tardif-Douglin, Indigo R. D. Miller, Stephanie C. Blitzer, David L. Gardner, Teresa M. Thompson, LaPonda Edmondson, David M. Levine

**Affiliations:** Ariadne Labs; Boston, MA, USA; CaroNova; Cary, NC, USA; North Carolina Healthcare Association; Cary, NC, USA; Scrum Inc; Cambridge, MA, USA; Division of General Internal Medicine and Primary Care, Brigham and Women’s Hospital; Boston, MA, USA; Harvard Medical School; Boston, MA, USA

**Author notes:** Corresponding Author: David Michael Levine Harvard Medical School, Brigham and Women’s Hospital, Division of General Internal Medicine and Primary Care 1620 Tremont Street, 3rd floor, Boston, MA 02120.

**Keywords:** Agile, Scrum, home hospital, hospital at home, intersectoral collaboration, diffusion of innovation, organization of work

## Abstract

**Background:** The diffusion of innovation in health care is sluggish. Evidence-based care models and interventions take years to reach patients. We believe the health care community could deliver innovation to the bedside faster if it followed other sectors by employing an organizational framework for efficiently accomplishing work. Home hospital is an example of sluggish diffusion. This model provides hospital-level care in a patient’s home instead of in a traditional hospital with equal or better outcomes. Home hospital uptake has steadily grown during the COVID-19 pandemic, yet barriers to launch remain for health care organizations, including access to expertise and implementation tools. The Home Hospital Early Adopters Accelerator was created to bring together a network of health care organizations to develop tools necessary for program implementation.

**Methods:** The Accelerator used the Agile framework known as Scrum to rapidly coordinate work across many different specialized skill sets and blend individuals who had no experience with one another into efficient teams. Its goal was to take 40 weeks to develop 20 “knowledge products,” or tools critical to the development of a home hospital program such as workflows, inclusion criteria, and protocols. We conducted a mixed methods evaluation of the Accelerator’s implementation, measuring teams’ productivity and experience.

**Results:** Eighteen health care organizations participated in the Accelerator to produce the expected 20 knowledge products in only 32 working weeks, a 20% reduction in time. Nearly all (97.4%) participants agreed or strongly agreed the Scrum teams worked well together, and 96.8% felt the teams produced a high-quality product. Participants consistently remarked that the Scrum team developed products much faster than their respective organizational teams. The Accelerator was not a panacea: it was challenging for some participants to become familiar with the Scrum framework and some participants struggled with balancing participation in the Accelerator with their job duties.

**Conclusions:** Implementation of an agile-based accelerator that joined disparate health care organizations into teams equipped to create knowledge products for home hospital proved both efficient and effective. We demonstrate that implementing an organizational framework to accomplish work is a valuable approach that may be transformative for the sector.

## Background

### Introduction

A large gap in health care exists between evidence-based knowledge and evidence-based practice,(1) often termed the “know-do gap.”(2) In 1962, Everett Rogers, a professor of rural sociology, published a seminal work on the diffusion of innovation.(3) He and others posited that the perception of the innovation, the adopters, and presiding contextual factors could impact the rate of spread.(4) In health care, as in other sectors, nearly all of these clusters often drive against rapid innovation. Yet by employing an Agile framework to organize work so that it is iterative, predictable, and joyful, other sectors have shown that rapid innovation is sustainable.(5) We hypothesized that applying similar organizational principles to the work of health care could facilitate the diffusion of rapid innovation.(6–9)

Hospitals are the standard site of care for acute illness in the US, but hospital care involves substantial risk and cost to the patient. One in 10 hospitalized patients will experience an adverse event, worse in older adults.(10–12) For decades literature has demonstrated that functional decline is a common adverse event associated with an acute hospitalization and is often related to the processes of care, rather than the patient’s underlying illness.(13) Despite evidence that movement improves patient outcomes, physical activity while hospitalized plummets.(14–17) As a result, 20% of formerly independent older medical patients require assistance walking after an acute hospitalization.(18) While admitted, 20% of older adults suffer delirium,(19–23) over 5% contract hospital-acquired infections(24), and many lose functional status that is never regained.(10,25–29) The cost of hospital care to the patient is the leading cause of personal bankruptcy.(16,29) Following hospitalization, many patients cycle among care settings, moving between rehabilitation and hospitalization and back again. (30,31)

Hospital-level acute care at home, or “home hospital,” was specifically designed to mitigate the risks associated with hospitalization and improve outcomes that are important to patients. Home hospital is the provision of hospital-level care in a patient’s home as a substitute for traditional inpastient hospital care.(32) Patients receive in-home nursing and physician care, intravenous medications, supplemental oxygen, laboratory and imaging diagnostics, and continuous vitals monitoring, among other services. (33) Importantly, home hospital is neither traditional “home care,” “home infusion,” nor “home hospice,” which often involve much lower acuity services such as twice weekly nursing and therapy. Internationally, nearly 40 years of evidence, including several randomized controlled trials, demonstrated that home hospital improves outcomes important to patients.(34–36)Home hospital patients have fewer 30-day readmissions, rate their experience of care more positively, feel more independent and less anxious, are more physically active, and maintain greater functional status.(33,34,37–42) From a clinical and administrative perspective, home hospital maintains patient safety and care quality, while reducing cost.(43,44)

Despite this evidence base, diffusion of the home hospital innovation has been slow, with fewer than 20 programs in existence for over a decade. The largest barrier was lack of a payment and regulatory pathway in the United States.

In November of 2020, the Centers for Medicare and Medicaid Services (CMS) issued the Acute Hospital Care at Home Waiver (AHCAH) in response to over-taxed hospital capacity created by the COVID-19 pandemic.(45) For the first time, the waiver created a nationwide regulatory and payment pathway for hospitals to deploy home hospital. The model has steadily grown from about a dozen programs prior to the pandemic to over 250 hospitals in 37 states.(46) This growth represents early adoption, yet a national survey of active programs revealed that program launch and growth remained challenging.(37) Barriers to implementation included clinician buy-in, appropriate patient identification, feasible workflows and protocols, and local content knowledge and expertise to guide the program. Many of these implementation barriers were so large that some programs that were able to secure a CMS home hospital waiver never enrolled patients.

Widespread success implementing home hospital during the CMS waiver is a rare policy window that could influence the fate of Medicare coverage for decades to come. Home hospital’s extensive, well-documented benefits to patients, caregivers, and hospitals and this time sensitive need for implementation support provided an opportunity to test a framework that accelerated the creation of tools necessary for home hospital.

### Launching the Home Hospital Early Adopters Accelerator

Our goal was to bring together health care organizations that had interest in starting or had recently launched a home hospital program to rapidly design and test knowledge products that could support program implementation. Knowledge products could include a patient admission workflow to admit a patient, patient-facing welcome packet, market scan of technology vendors, or list of requirements for commercial reimbursement contracts. We coupled our experience with home hospital and survey data from participating organizations to identify 20 knowledge products that would create a substantial portion of the tools needed to deploy a program. To make high quality tools available as soon as possible, we planned to create those 20 knowledge products in 40 weeks. We also planned to study the team’s performance.

### Using Scrum to accelerate home hospital tool creation

Extensive knowledge of the home hospital model is often not common among health care organizations. To effectively deploy a program like home hospital, a health care organization charged with implementation needs access to deep expertise and experience, rather than a broad implementation guide or the abundance of available published literature. Traditional, one-on-one technical assistance approaches risk personnel-heavy, expensive, slow hospital-by-hospital diffusion of innovation.(47,48) We searched for a methodology to bring together a network of health care organizations to accelerate their knowledge and expertise of home hospital. We gravitated toward Agile methodologies given their success in other sectors and their ability to break complex projects into manageable chunks of work.(49) They have become more common in health care to rapidly organize teams, fulfill diverse needs faster, offer end-users involvement in the development process, all while using fewer resources and improving quality.(7,50–54)

Of the Agile methodologies, we chose to use Scrum due to its flexibility, low barrier to entry (e.g., freely available tools),(55,56) industry evidence that it accelerates work,(51,57–59) and prior experience among our own team members. Scrum employs an iterative and incremental approach to optimize predictability, control risk, and make work joyful. The members of a Scrum team collectively have all the necessary skills and expertise to do the work or can easily acquire such skills as needed. Scrum creates the structure for people to come together regardless of their background or area of expertise.(55) Scrum began as a tool for organizing software development teams, but it has been adopted across industries.(55,57–64) Scrum has been used in health care to design electronic health record tools,(65) develop communication tools for cancer patients,(66) and develop a telemedicine care pilot.(67) We hypothesized Scrum could catalyze coordination across disparate health care organizations to rapidly build the tools necessary for a home hospital program to launch, instead of the often years-long pursuance of developing workflows, technology stacks, and protocols.(57,58,63) We also expected that the Accelerator would break down the traditional siloed development that occurs among health care organizations.

The Scrum framework is purposefully limited to a few essential elements (**Additional file 1)** so the people using it can shape their work to best fit their skills, relationships, and interactions. It involves 3 roles, 5 events, and 3 artifacts (**Figure 1**). The team roles include the Product Owner who has deep knowledge of the field and user requirements, the Scrum Master who leads and organizes the team toward its products, and Developers who have the knowledge to develop the team’s products. The Scrum process has several steps **(Figure 2)**:

**Figure 1.**
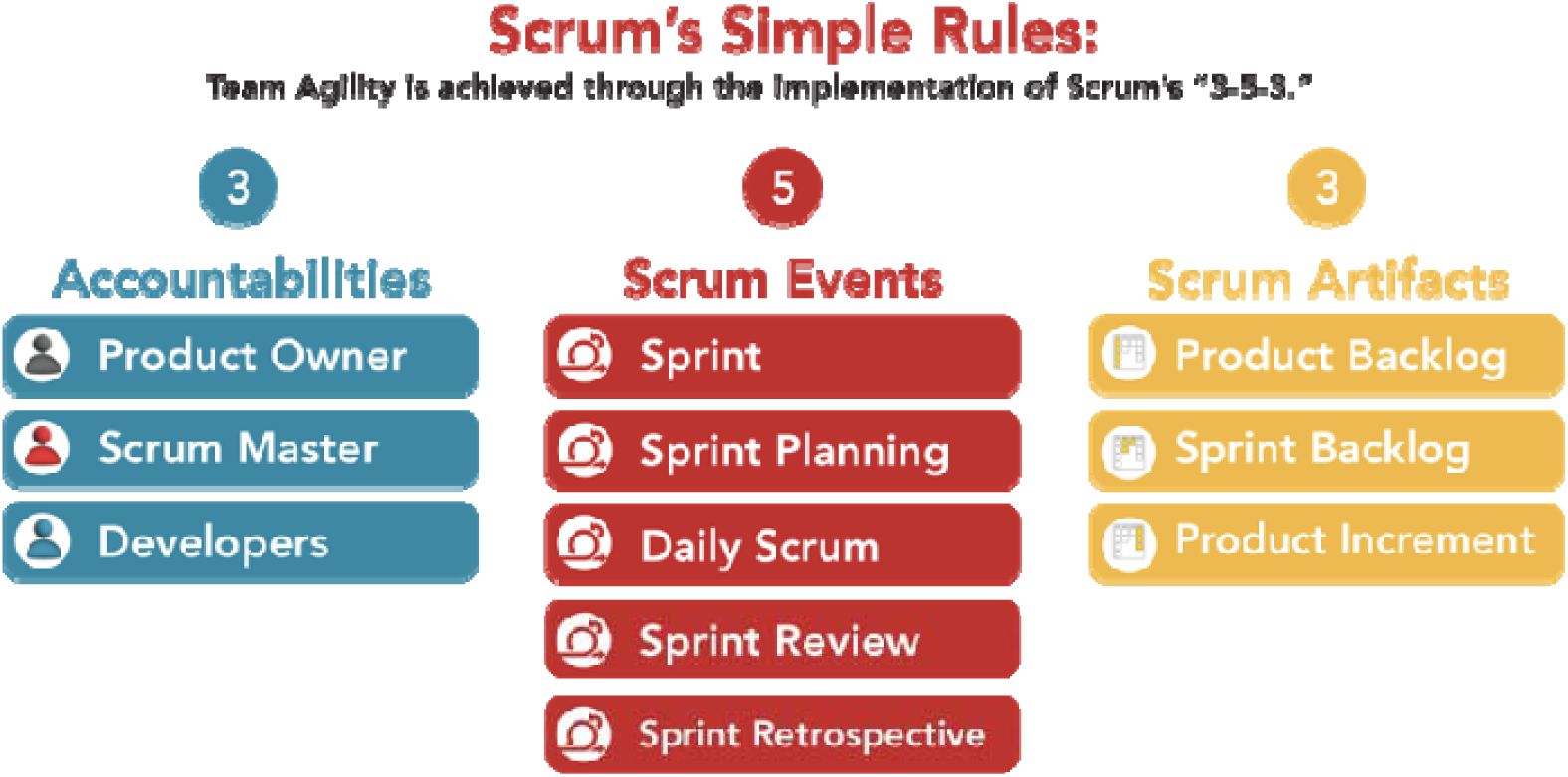
Scrum Implementation. Refer to Table 1 for details on each Scrum element

**Figure 2.**
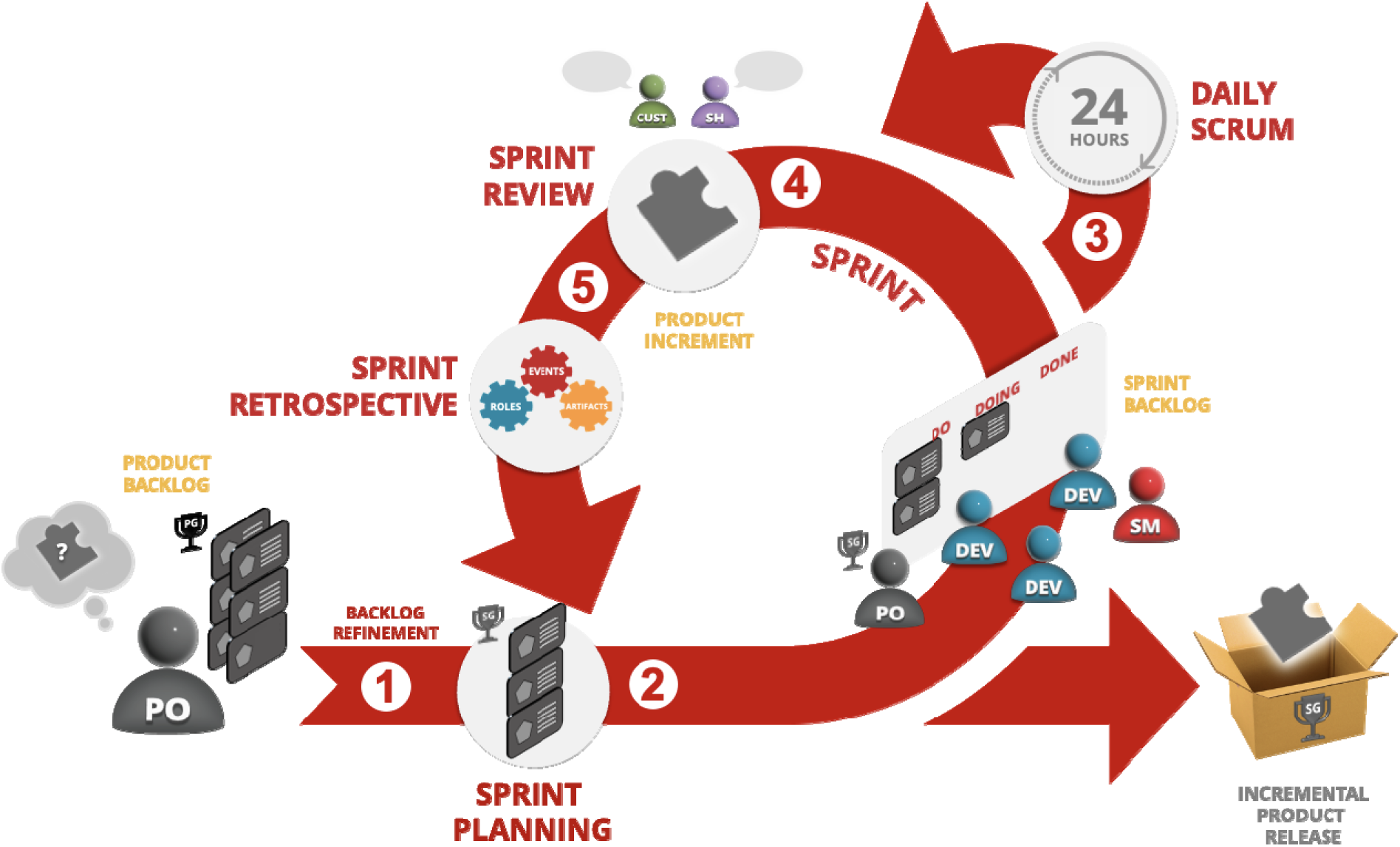
The Anatomy of a Sprint. Refer to the Scrum Glossary in Additional file 1 for details on each Scrum element

**Table 1.** Scrum elements and adaptations made for the Accelerator.

| Scrum Element | Typical Scrum | Accelerator Adaptations<sup>a</sup> |
| --- | --- | --- |
| **Scrum Roles** |  |  |
| Product Owner (PO) | Full-time<br>Part of backlog planning and the sprint review.<br>Defines and communicates the product goals<br>Determines and prioritizes Product Backlog items | Part-time |
| Scrum Master | Full-time<br>Leads Sprint Planning, Daily Scrum, Sprint Review, and the Sprint Retrospective<br>Helps the Developer team reach peak performance by supporting their ability to collaborate effectively | Part-time<br>Trains Developers to use the virtual collaboration tools<br>Coordinator support for whiteboard updates, managing digital access, troubleshooting digital issues, and scheduling |
| Developer(s) | Full-time<br>Use their skills to accomplish the work needed to complete each User Story<br>Expected to attend all Scrum events (below) | Part-time, often while managing clinical responsibilities |
| **Scrum Events** |  |  |
| Sprint | Timing: 2-week Sprints<br>Participants: Developers, Product Owner, and Scrum Master.<br>Goal: create a product increment. | Timing: 1-week Sprints (2 Sprints per knowledge product)<br>Participants: +coordinator<br>Goal: no change |
| Sprint Planning | Timing: 2-4 hours (backlog refinement ~2 hours)<br>Participants: as above<br>Goal: refine the product backlog ahead of the sprint, elect backlog items and prepare to accomplish them in the | Timing: 2.5 hours (backlog refinement 1 hour every other week)<br>Participants: as above + 1-2 Developers<br>Goal: no change |
|  | upcoming sprint |  |
| Daily Scrum | Timing: 15 minutes daily<br>Participants: Developers, Product Owner, and Scrum Master<br>Goal: find and remove impediments to Developer progress | Timing: no change<br>Participants: +Product Owner + coordinator<br>Goal: no change |
| Sprint Review | Timing: once at the end of every Sprint for four hours or less.<br>Participants: Developers, Product Owner, and Scrum Master.<br>Goal: acquire product owner feedback on whether the product increment meets requirements | Timing: 1-hour<br>Participants: +coordinator, +stakeholders<br>Goal: acquire stakeholder feedback on whether the product increment meets requirements |
| Sprint Retrospective | Timing: Once at the end of every Sprint for three hours or less<br>Participants: Developers, Product Owner, and Scrum master<br>Goal: identify what went well, what could have gone better, and what change (Kaizen) to test in the next sprint | Timing: 1-hour<br>Participants: +coordinator<br>Goal: no change |
| **Scrum Artifacts** |  |  |
| Product Backlog | Developed and maintained by the PO. Commonly uses automated project management tools<br>Each user story has an estimated number of story points, based on the level of effort required as determined by consensus among the Developers | Developed and maintained by the PO with survey input from the Accelerator sites and support from the coordinator |
| Sprint Backlog | Developed and maintained by the Scrum Master and Developers. Commonly uses automated project management tools | Participants: +coordinator |
| Product Increment | Each Sprint typically produces a product increment, for which there may be multiple user stories required. A product increment meets basic functionality for use in a real-world setting. |  |

<sup>a</sup>: only differences are noted

#### Step 1: Backlog Refinement

The Scrum process begins with Backlog Refinement (step 1), where the Scrum team defines the various products and breaks them into well-defined groups of tasks and minimum requirements needed to create a product that is ready to use (referred to as a Product Increment). To accomplish the Product Increment, the Scrum team creates multiple User Stories that represent chunks of work. These User Stories form the Product Backlog.

#### Step 2: Sprint Planning

At Sprint Planning (step 2), the Developers select User Stories from the Product Backlog to include in the Sprint. Developers further refine the User Stories and assign team members to the tasks required to complete the User Story. These tasks form the Sprint Backlog. Then the Scrum team estimates the level of effort required to accomplish the User Stories using Story Points (a relative measure of effort described below) and commits to the Sprint.

#### Step 3: Daily Scrum

Each day the team holds a Daily Scrum (step 3), where each member notes what items from the Sprint Backlog they accomplished, their impediments, and what they will work on that day.

#### Step 4: Sprint Review

If the team believes the Sprint successfully achieved its goal, the team holds a Sprint Review (step 4), where they present a ready-to-use product to the project’s stakeholders (critical reviewers of the work) for feedback on how well the product meets minimum requirements for real-world use.

#### Step 5: Sprint Retrospective

The team holds a Sprint Retrospective (step 5) to identify successes and areas for improvement. The team then commits to an improvement by doing one thing differently in the next Sprint.

## Methods

### Recruitment of participants

To solicit participants, we created a public request for participation in the Home Hospital Early Adopters Accelerator (referred subsequently as “the Accelerator”). Any health care organization was welcome to participate, including hospitals, primary care offices, home health agencies, and health care insurers. As a partnership between Ariadne Labs (an initiative of Harvard T.H. Chan School of Public Health and Brigham and Women’s Hospital) and CaroNova (an initiative of The Duke Endowment, the North Carolina Healthcare Association, and the South Carolina Hospital Association), the Accelerator was available to sites in North and South Carolina free of charge, while sites elsewhere paid a small participation fee. We asked sites to apply by describing their plans for launching a home hospital program or to detail their current program. We also required a commitment from executive leadership that the health care organization would pledge to send staff to help develop at least 5 knowledge products over the 40 weeks. We accepted all sites that submitted a complete application.

### Adapting Scrum for the Accelerator

As is common when using Scrum, we needed to make adaptations to the traditional Scrum processes to fit our goal due to the multicenter nature of the work and the Accelerator’s constantly changing teams **(Table 1).** Typically, a Scrum team works together full time for months or years on end. Instead, we asked participating health care organizations to staff each Sprint with Developers who had content expertise in the planned knowledge product. Therefore, the Developers were not longstanding colleagues but were instead thrust together for a maximum of 2 weeks without prior knowledge of Scrum or experience working with one-another. Only the Scrum Master, Product Owner, and supporting project coordinator remained consistent from Sprint to Sprint (**Table 2**).

**Table 2.** The home hospital early adopters accelerator team.

| Team member | Description of the Accelerator Role | Scrum Events attended |
| --- | --- | --- |
| Product Owner | Established requirements for a knowledge product<br>Led the prioritization of Sprint and Product Backlog items | All |
| Scrum Master | Ensured adherence to core Scrum principles | All |
| Project Coordination | Coordinated assignment of Developers to Sprints<br>Scheduled all meetings and emailed key updates<br>Sent assessment surveys | All |
| Developers<sup>a</sup> | Brought clinical and health care management experience<br>Executed the tasks needed to complete each Sprint | All |
| Stakeholders<sup>a</sup> | Brought clinical and health care management experience<br>Provided feedback on the knowledge products during Sprint Reviews | Sprint Review |
| Consultants<sup>b</sup> | Provided ad hoc input on knowledge products in their field of expertise or with skillsets the Developers did not have | None |
| Subject Matter Experts<sup>b</sup> | Individuals with home hospital expertise who could answer Developer questions | None |

<sup>a</sup>: Developers and stakeholders participated when they signed up to work on the development of a specific knowledge product (Developers) or review and provide feedback on a knowledge product (stakeholders).
<sup>b</sup>: Consultants and subject matter experts participated when contacted for input on Knowledge products

In addition, all Scrum activities were performed remotely. It was unclear at the start of the Accelerator whether we could achieve sufficient teamwork and assimilate team members to Scrum processes in such a short period of time. Because of the rotating Developers, we asked a small cohort of Accelerator participants to support our core team in producing the Product Backlog, instead of involving the Developers. This enabled us to give the newly forming Scrum teams a head start with highly developed User Stories that usually required only minor refinement and clarification during each Sprint.

To run this fast-paced, distributed Scrum adaptation, we turned to a package of synchronous and asynchronous collaboration tools to connect team members. We used standard software tools like Zoom, Dropbox, and Microsoft Office. To create a digital whiteboard and collaboration space, we used Miro. Our Miro board housed content for our Sprint Backlog, Sprint Planning, Daily Scrum, and Sprint Retrospective (**Figure 3**). It also served as a knowledge management platform housing a central repository for links and prior knowledge products. Developers created shared norms around collaboration for each Sprint.

**Figure 3.**
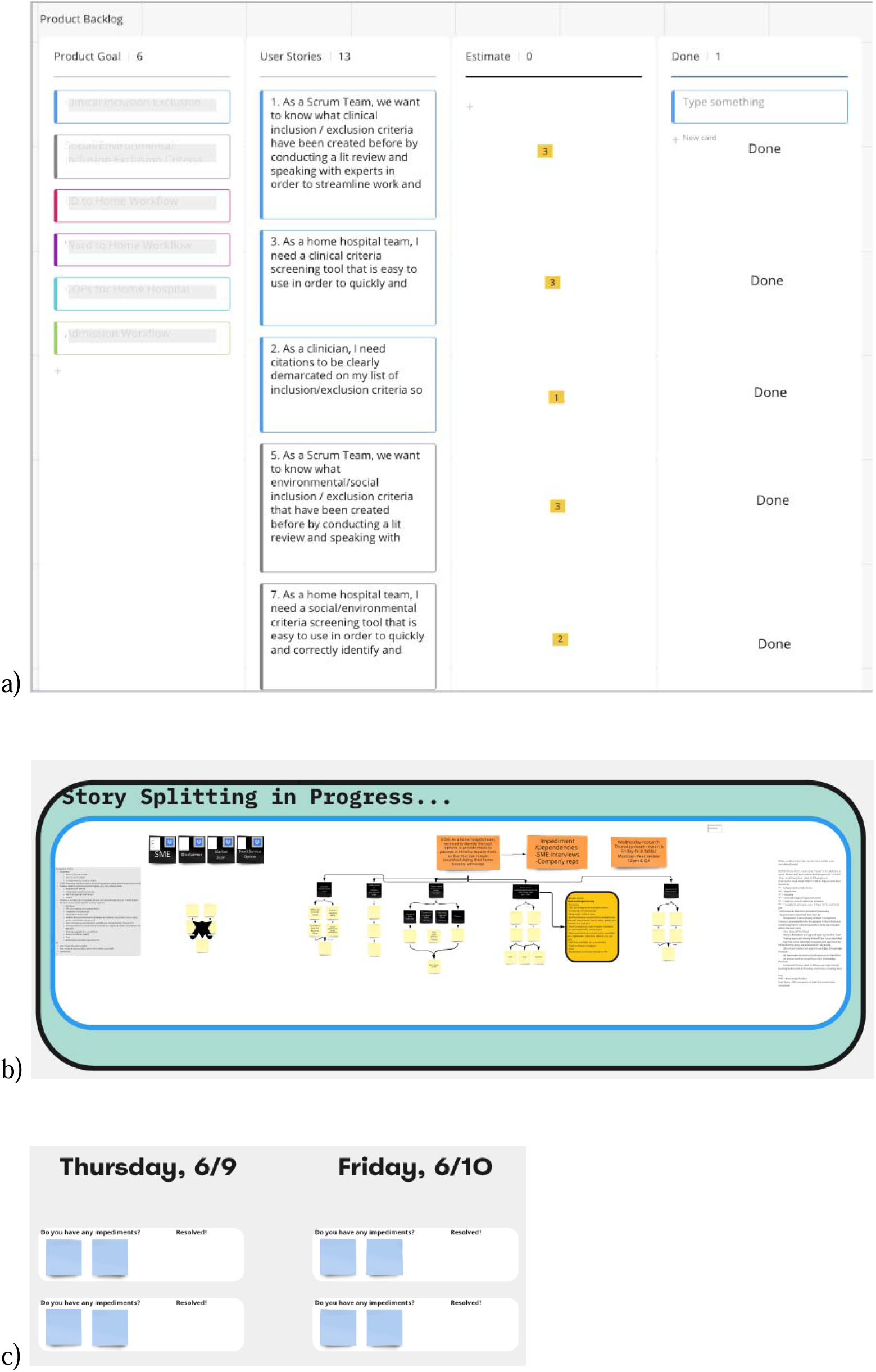
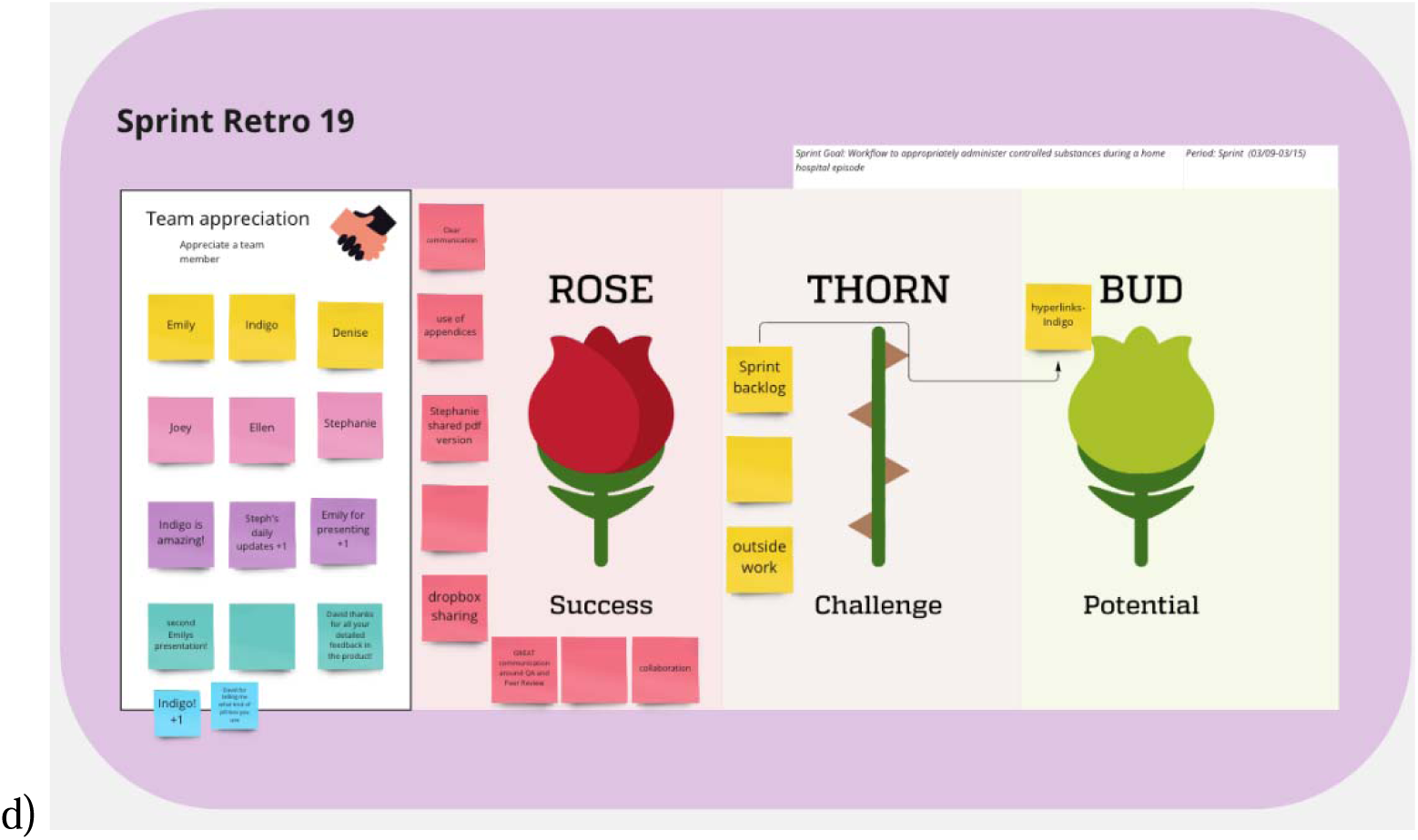
Remote Scrum via virtual whiteboard. a) Sprint Backlog; b) Sprint Planning; c) Daily Scrum; and d) Sprint Retrospective.

During launch preparation, we required role-based training (**Figure 4**). The team from CaroNova and Ariadne took the Scrum Startup for Teams training that taught the basics of Scrum (9 hours). Scrum Inc. additionally provided the team a coach with whom we could consult as needed for Scrum methodology questions (10 hours of coaching time). To train the incoming Developers from individual sites, we asked them to review two articles and a video about Scrum and home hospital (1 hour). We also asked Developers to test their access to the tools ahead of the Sprint Planning and offered office hours with the Scrum Master and project coordinator to resolve any remaining technological or knowledge barriers to Sprint participation.

**Figure 4.**
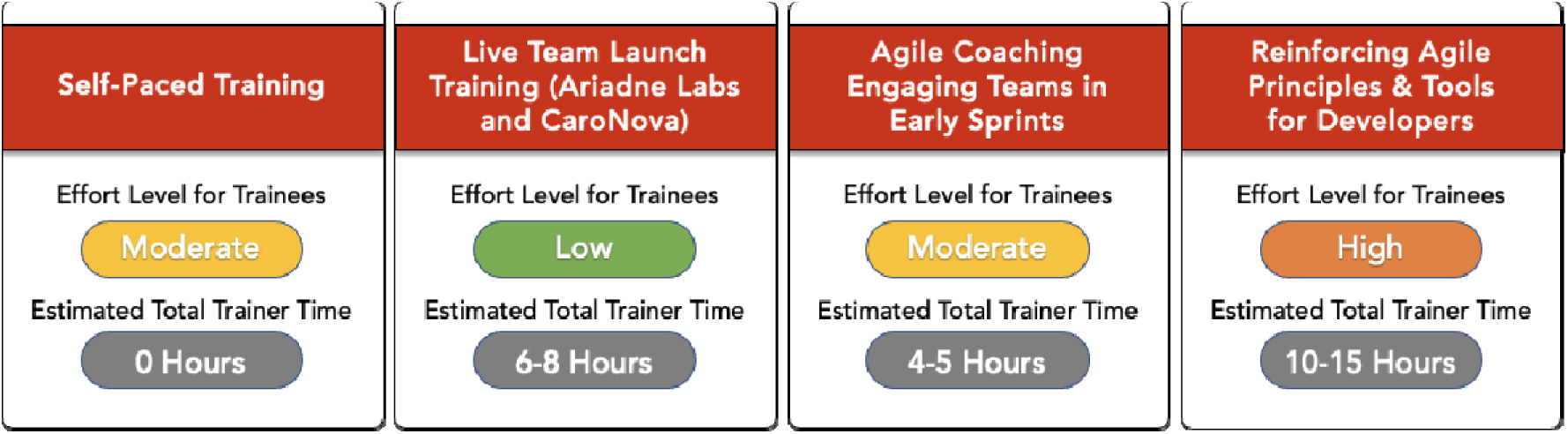
Training type and level of effort.

### Knowledge product creation

For Developers, weekly Sprints began with Sprint Planning, which included estimating the level of effort required to accomplish the selected backlog items. Level of effort is quantified as “Story Points” instead of an estimate of time required for each item. Story Points represent the level of complexity and novelty of the tasks within the ensuing Sprint, with more points equating to more work. Velocity represents the Story Points per sprint. A Scrum Master calculates the planned Velocity at the beginning of the Sprint and the actual Velocity at the end of a Sprint. High-functioning teams may therefore exceed initial Scrum Velocity estimates if they are able to accomplish more work during a Sprint. As an example, a team may plan to accomplish 2 User Stories as part of a 1-week Sprint. If each User Story is estimated at 3 Story Points, the planned Velocity is 6 Story Points. If the team is very efficient and decides to include another User Story worth 2 Story Points, their actual Velocity is 8 Story Points.

### Evaluation of the Accelerator

At the end of each Sprint, we collected developer feedback with an electronic survey using REDCap (Research Electronic Data Capture, Vanderbilt University, Nashville, TN, a fully HIPAA-compliant web application). The Developer survey measured participant experience with the process of the Accelerator, and included items related to one’s experience participating in the Sprint (Likert scale: 1, strongly disagree; 5, strongly agree) (**Additional file 1)**. For example, to assess experience we asked, “The Scrum team worked well together” and “It was difficult to create the knowledge product created during this Sprint.” There could be multiple survey responses per Scrum participant because a Developer who participated in two knowledge products would complete the survey twice.

At the end of the Accelerator, we conducted semi-structured interviews with Developers and stakeholders to gather further depth and context on their experience as well as collect feedback on how the Accelerator format could be improved (**Additional file 1)**. A member of the study team (MPD; female, MPH) conducted each interview with the participant over an encrypted connection. We audio recorded and transcribed the interviews for analysis using Dedoose qualitative software. A multidisciplinary research team with home hospital, agile methodology, and qualitative research expertise developed an initial codebook deductively based on the evaluation aims and interview guide questions. A member of the study team (MPD) used an inductive coding approach, adding emerging themes or codes throughout the process. One coder (MPD) performed a thematic analysis of transcriptions, and a second coder (SB; female, BA) double coded a subset of the interviews. Any disagreements were first discussed between the two coders to assess and improve intercoder agreement. (68–70) If needed, the coders achieved consensus on any remaining disagreement through discussion with a third researcher (DML; male, MD).

## Results

### Participants and sites

Of the eighteen Accelerator sites, most were large (62.50%), urban (83.33%) hospitals and hospital systems (**Table 3**). Less than half (44.44 %) of the sites had begun enrolling home hospital patients as of September 2021 when the Accelerator launched.

**Table 3:** Site characteristics.

| Characteristic<sup>a</sup> | Sites (n=18) |
| --- | --- |
| Geography, n (%) |  |
| Rural | 3 (16.67%) |
| Urban | 15 (83.33%) |
| Organization type, n (%) |  |
| Hospital | 16 (88.89%) |
| Home health agency | 1 (5.55%) |
| Payor | 1 (5.55%) |
| Total population of towns or cities served, n (%)<sup>b</sup> |  |
| <100,000 | 6 (33.33%) |
| 100,000-399,999 | 6 (33.33%) |
| 400,000+ | 6 (33.33%) |
| Bed size, n (%)<sup>c</sup> |  |
| 1-299 | 5 (31.25%) |
| 300-499 | 1 (6.25%) |
| 500+ | 10 (62.50%) |
| Enrolling home hospital patients at Accelerator start, n (%) | 8 (44.44%) |

<sup>a</sup>: We based site characteristics on individual facilities unless Developers and stakeholders from the site represented multiple facilities within a system. In such instances, we based site characteristics on the entire health care system.
<sup>b</sup>: For the total population of towns or cities served, this is either the population of the local city for single hospital facilities, the combined population of the cities of each of the hospital facilities for systems, or the population of the city or cities a home health agency or payor serves.
<sup>c</sup>: Hospitals only.

There were 61 Developers and 44 stakeholders. Overall, most Developers were white (83.61%) and many were hospital administrators (42.62%), followed by nurses (24.59%) (**Table 4**). Most stakeholders also were white (88.64%) and most were hospital administrators (38.64%), followed by nurses (20.45%). Physicians accounted for 11.48% of developers and 18.18% of stakeholders.

**Table 4.**
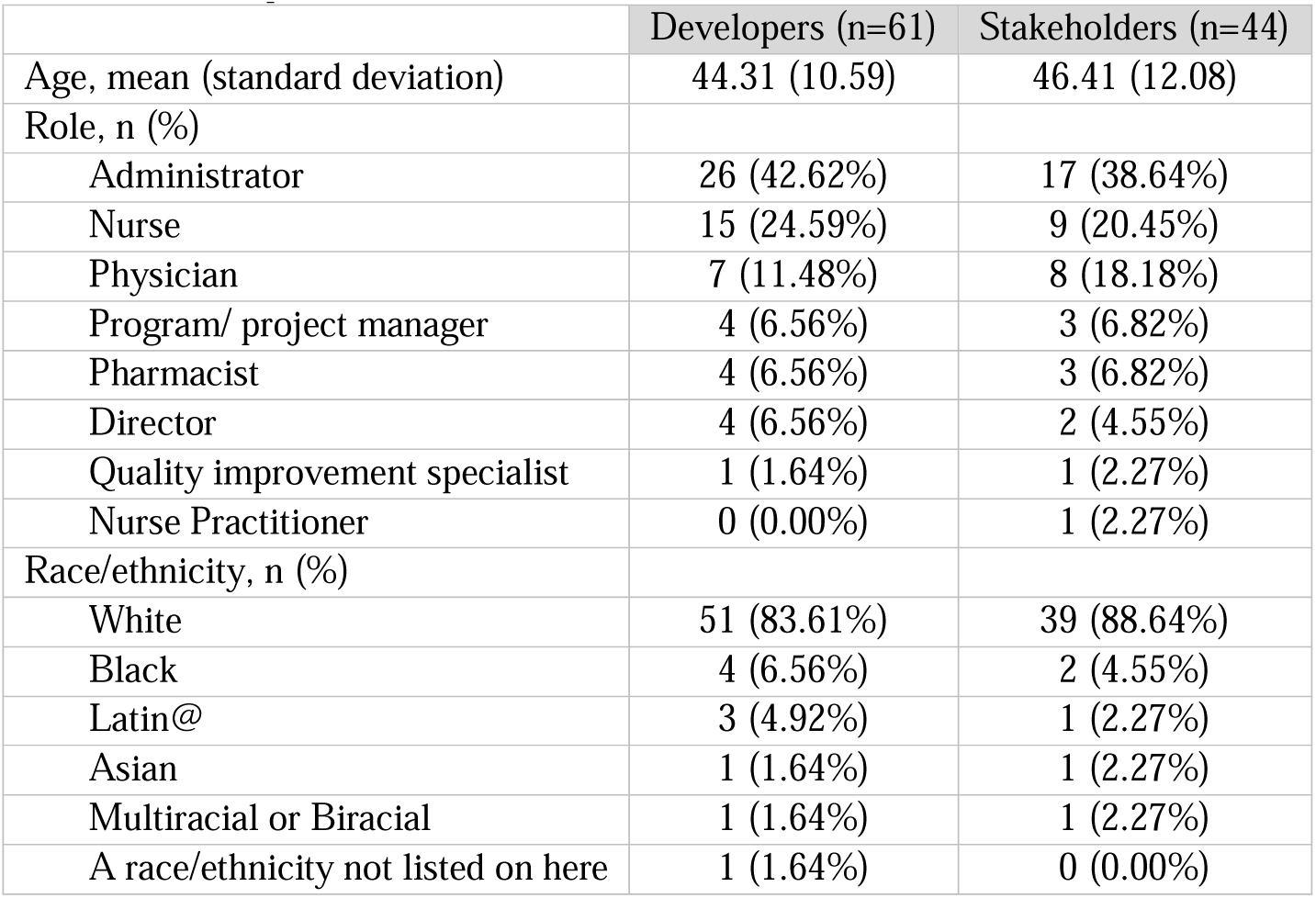
Participant characteristics.

### Knowledge product creation metrics

The mean planned Velocity was 2.4 Story Points per Sprint (**Figure 5**). The mean actual Velocity was 3.0 Story Points per Sprint. This ultimately led to completing the 20 knowledge products 8 sprints early for a 20% reduction in schedule.

**Figure 5.**
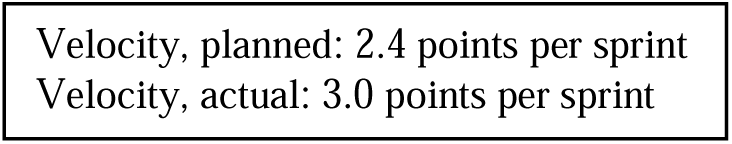
Product burndown and performance velocity.

### Participant and site experience

#### Quantitative findings

The results of this survey suggest Developers were broadly satisfied with the process and the output of the Accelerator **(Table 5).** Developers reported working approximately 5.65 hours (SD, 4.03) hours each week. Most believed the Scrum team worked well together (40.4% agreed, 57.1% strongly agreed), and the Sprints were productive (38.8% agreed, 59.6% strongly agreed). The majority of respondents also thought the Scrum team spent an acceptable amount of time working on each knowledge product (46.8% agreed, 48.1% strongly agreed). Most developers and stakeholders were pleased with the results, agreeing that the Sprint produced a high-quality product (32.7% agreed, 64.1% strongly agreed).

**Table 5:** Developer experience.

|  | (n=156) |  |  |  |  |
| --- | --- | --- | --- | --- | --- |
| The scrum team... | Strongly disagree (%) | Disagree (%) | Neutral (%) | Agree (%) | Strongly agree (%) |
| ...worked well together | 1 (0.6) | 2 (1.3) | 1 (0.6) | 63 (40.4) | 89 (57.1) |
| ...was productive | 1 (0.6) | 0 (0) | 3 (1.9) | 59 (38.8) | 93 (59.6) |
| ...spent an acceptable amount of time working on the product | 1 (0.6) | 1 (0.6) | 6 (3.8) | 73 (46.8) | 75 (48.1) |
| ...produced a high-quality product (f4) | 1 (0.6) | 0 (0) | 4 (2.6) | 51 (32.7) | 100 (64.1) |

#### Qualitative findings

From the qualitative analysis of Developer and stakeholder interviews, we identified several benefits of participating in the Accelerator including collaboration and knowledge sharing (**Table 6**). We also found multiple perceived advantages of utilizing the Scrum framework as well as some challenges.

**Table 6.**
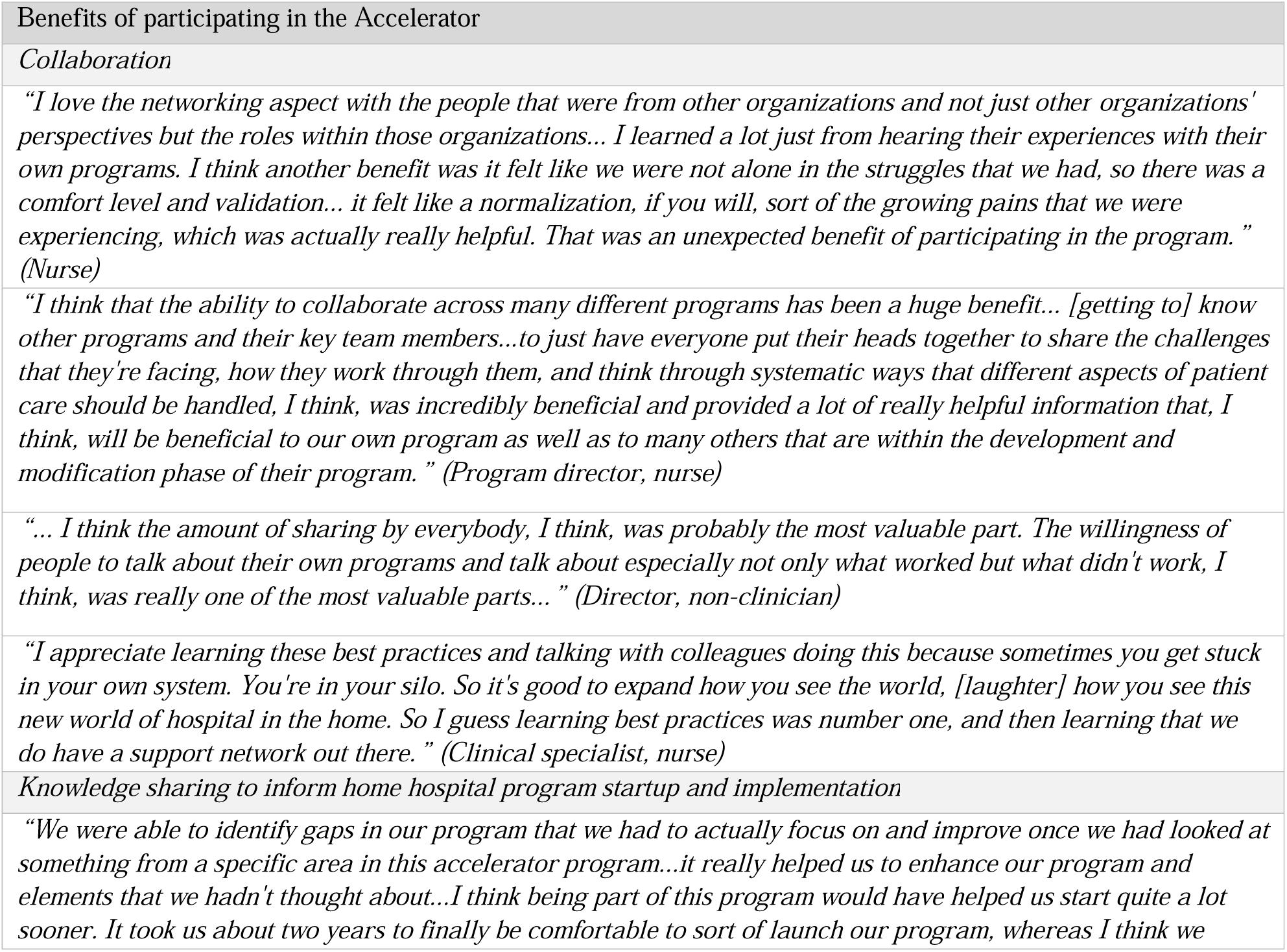
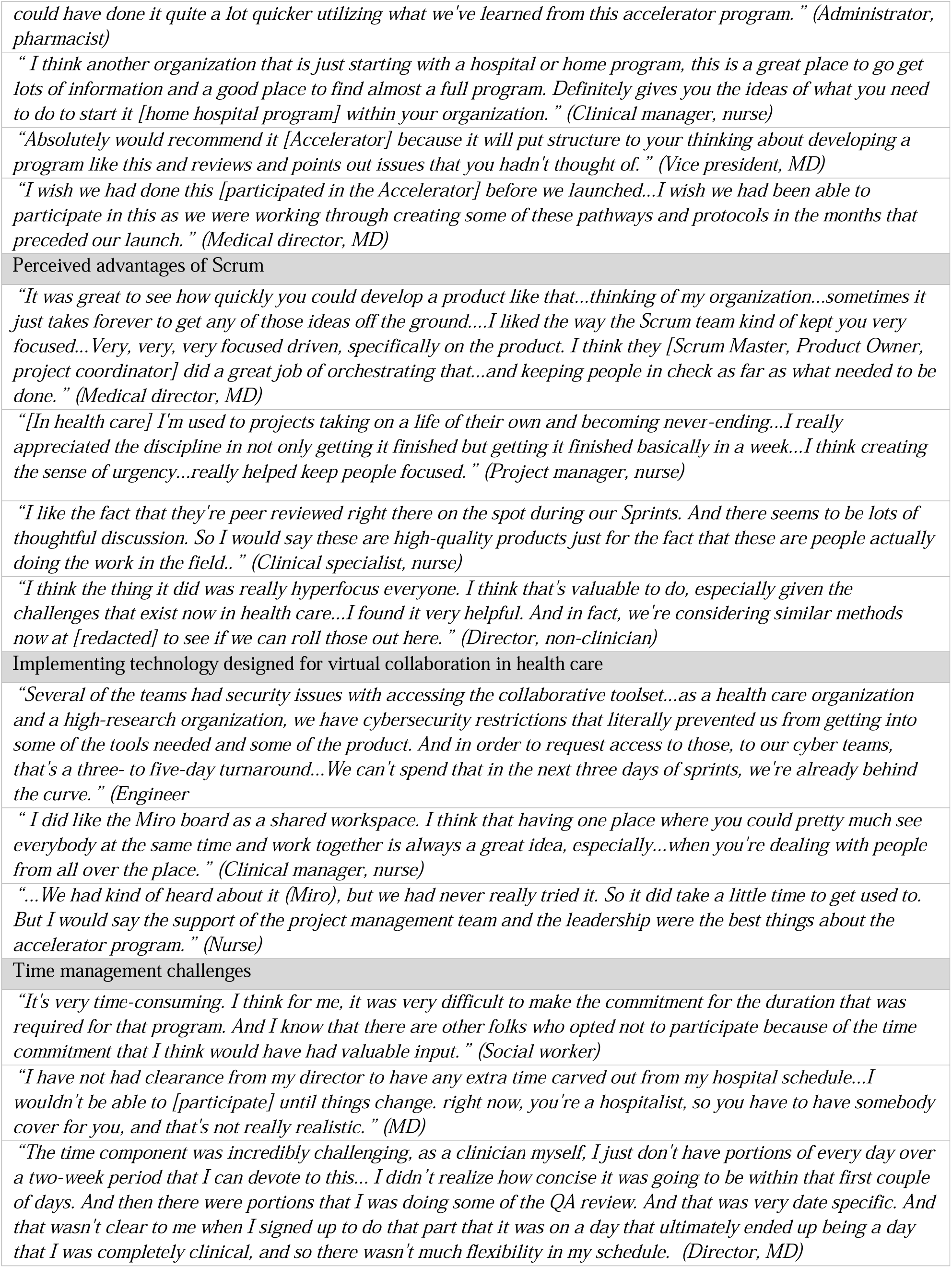
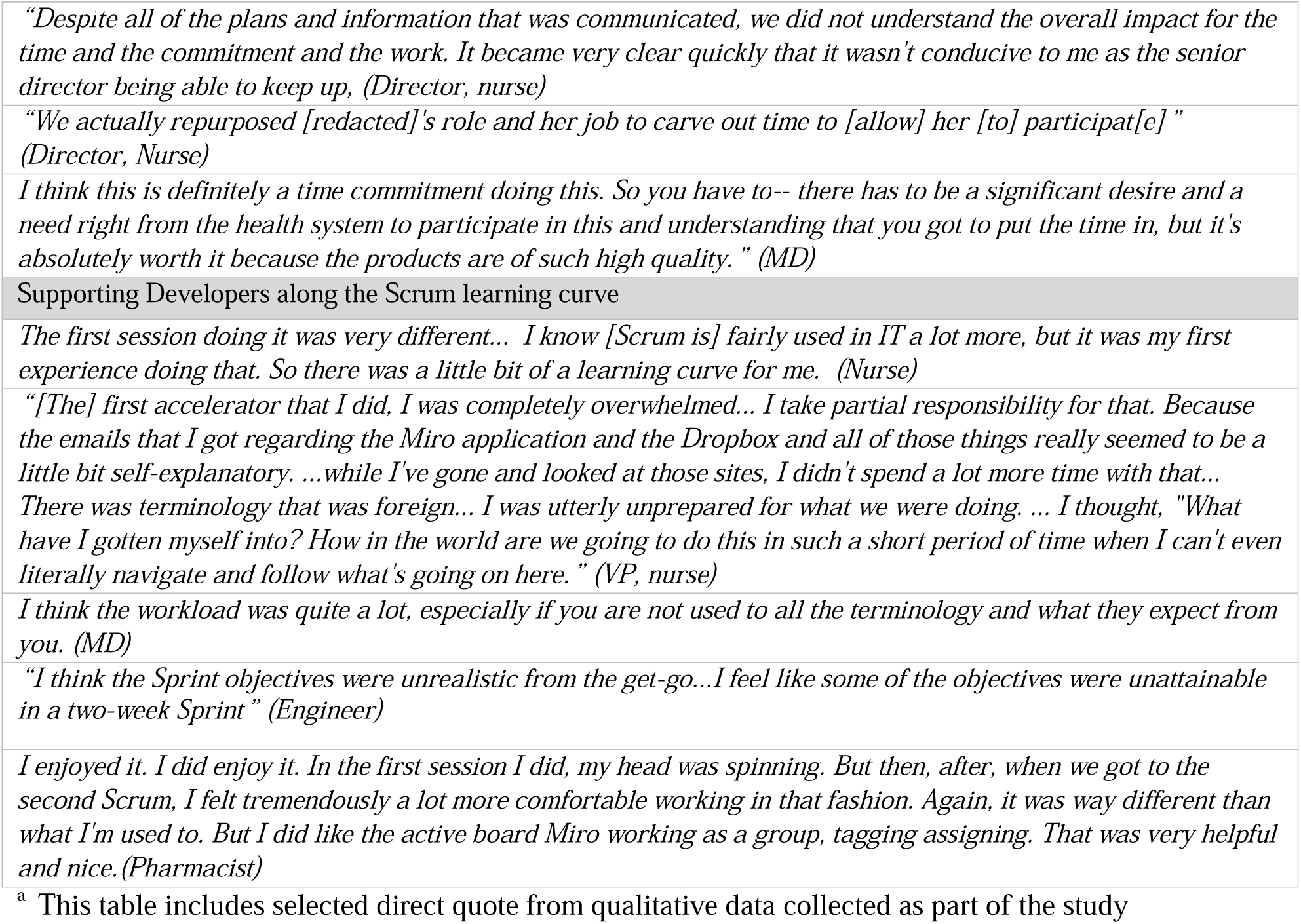
Qualitative findings of Developers’ experience ^a^.

Accelerator participants spoke at length about the benefits of collaborating within the Accelerator’s Scrum framework. They saw value that it was not only with other home hospital programs, but with other individuals who had diverse backgrounds and expertise. Participants appreciated the opportunity to network with and learn from other home hospital programs by discussing challenges, sharing and receiving feedback on new ideas, and hearing about the experiences of other participants.

Another perceived benefit of participation was the ability to learn firsthand how a health care organization can launch their own home hospital program. Participants from organizations that had an existing home hospital program cited the benefits of crosschecking their existing tools and resources, like clinical and administrative workflows, with those being developed in the accelerator. This helped identify gaps in existing programs and inform improvements.

Through their experience with the Accelerator, participants noted that the Scrum framework was a positive change from their standard way of working on projects in health care. Scrum helped focus and organize participants, it made the process of creating knowledge products efficient (without sacrificing quality), and the meeting format allowed for more collaboration. Overall participants said that more work was accomplished than thought possible.

The Scrum framework is highly collaborative and adapting Scrum to a virtual environment requires use of virtual collaboration tools. Some Developers’ organizational policies and firewalls made it hard to access shared documents **(Table 6)**. This required workarounds, such as sharing the latest version of documents by email or designating someone with access to upload documents on behalf of a Developer without access. When Developers could access virtual collaboration tools, they were not universally familiar with them. The program coordinator became adept at helping Developers quickly log into and navigate both the virtual whiteboard and the shared cloud drive. Our model repeatedly brought together new, multidisciplinary groups of developers to work on each knowledge product. Matching the right individuals with the right expertise for a given knowledge product was vital but could be challenging. We provided sites with information about the required expertise for each knowledge product far in advance of Sprints so they could identify Developers. However, some Developers were not prepared for the hours of work required for each Sprint and found it hard to balance their other work priorities (**Tables 6**). Occasionally, Developers felt their expertise was poorly matched to the knowledge product.

In most cases, Developers were working on a Scrum team for the first time. Some felt overwhelmed by the unfamiliar Scrum terminology and the use of a virtual white board for live collaboration during the initial Sprint Review (**Table 6**). Others found the format of the Sprint to be very fast paced or with unnecessary components. This was a manageable learning curve, however, as Developers consistently expressed that their second Sprint was much easier than their first.

## Discussion

Implementation of an Agile-based accelerator that brought together 18 previously unconnected health care organizations was feasible, acceptable, and efficient in generating health care products faster than anticipated, requiring 20% less time. The Accelerator successfully produced its desired 20 knowledge products ahead of schedule using an adapted Scrum framework. We believe organizational frameworks like Scrum are underutilized in health care as a method to drive collaboration and develop knowledge products to address various health care problems. Below we discuss the challenges we faced, lessons learned, and advice for future Accelerators. We describe our overarching recommendations and the steps required to launch an Accelerator in **Figure 6**.

**Figure 6.**
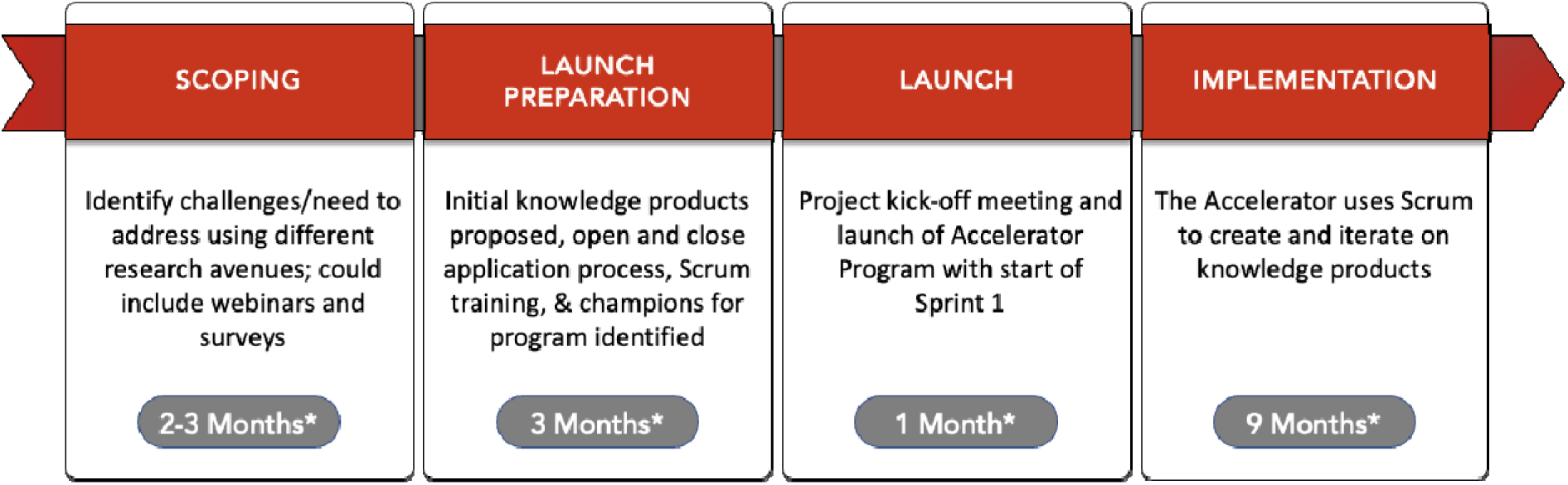
Steps to launch a Scrum-based accelerator. *Months noted reflect the timeframe for scope to implementation of the Accelerator. This timeline may be different for other programs.

### Implementing technology designed for virtual collaboration in health care

We recommend assessing Developers’ comfort with tools before a Sprint to facilitate targeted support. Consider holding early conversations with site IT staff to ensure needed access. Offering office hours for ongoing training and dedicated technical support helps further reduce barriers.

### Recruiting engaged Scrum team members

We recommend having backup Developers that the project coordinator can enlist should others have to bow out or be a poor fit for the knowledge product. We asked Developers who might have to miss a Daily Scrum meeting due to a competing conflict to use the digital whiteboard asynchronously to inform the rest of the team of their progress or any impediments. We also provided several subject matter experts who could be brought in quickly when an area of expertise was lacking on the Developer team. To make the time commitment explicit, we further recommend estimating for Developers the total weekly number of hours they should reserve on their calendar for Scrum participation.

### Supporting Developers along the Scrum learning curve

To acclimatize Developers to the Scrum framework, the Scrum Master included a 5-minute Scrum overview at the beginning of each Sprint Planning, as well as weekly office hours. To adjust expectations for new Developers, we also consistently communicated that after one Sprint, the learning curve eased. We recommend clear training requirements for Developers in preparation for Sprint participation. This could include mandatory training session attendance or use of learning management systems to ensure asynchronous completion of learning content.

This work has limitations. While we did have one international healthcare organization participate in the Accelerator, most of our participating sites were U.S. based organizations therefore limiting generalizability. In addition, most of our participating sites were large, urban hospitals. Further testing is required to see if smaller and rural organizations would have the capacity to participate in an Accelerator of this nature. In addition, an adapted Scrum framework worked for our Accelerator, but it remains to be seen if a similar method can be successfully implemented in other sectors of healthcare.

## Conclusion

Health care has an innovation dilemma: innovative existing tools that need rapid deployment and implementation in weeks to months arrive at the bedside often years later. We believe this dilemma is partly due to how we structure quality improvement and implementation efforts in health care. Organizing team members via Scrum facilitated productivity rarely seen inside health care while maintaining high quality products.

We feel this framework could be easily applied to many of health care’s complex and simple problems and provide transformative speed and accuracy to a team’s efforts. For example, hospitals could use Scrum to develop a readmission reduction program, a new surgical enhanced recovery after surgery pathway, or a discharge before noon process. Primary care could develop ideal cancer screening workflows, chronic disease management pathways, or urgent care triage protocols. Researchers could use Scrum to optimize their machine learning model, to find chemical compounds more efficiently, or to optimize deployment of a trial protocol in the field. To our knowledge, others in health care have rarely published metrics such as productivity velocity or the methods to organize busy health care organizations. We view this framework of organizing work as something missing from most health care efforts. Changing the organizational principles by which health care practitioners work could counter the prevailing trends that prevent the rapid diffusion of innovation and lead to a transformative closure of the know-do gap for millions of patients.

## Data Availability

The datasets generated and/or analyzed during the current study are not publicly available due to individual privacy concerns but are available from the corresponding author upon reasonable request.

## List of Abbreviations

CMS: Centers for Medicare and Medicaid Services
Dev: Developer
PO: Product Owner
REDCap: Research Electronic Data Capture
SD: Standard Deviation
SM: Scrum Master

## Declarations

## Ethics approval and consent to participate

This study was granted exemption by the Mass General Brigham IRB (2021P002411). All methods were carried out in accordance with relevant guidelines and regulations. Informed consent was obtained from all subjects.

## Consent for publication

Not applicable

## Authors Contributions

MD had full access to all the data in the study and takes responsibility for the integrity of the data and the accuracy of the data analysis. SB carried out administrative, technical, or material support. MD and MTD conducted statistical analyses. MD, MTD, SB, TT, DG, and DL drafted the manuscript. The study was supervised by DL. All authors contributed to study concept and design, interpretation of data and critical review of the manuscript for important intellectual content.

## Acknowledgements

The authors acknowledge Matthew Jacobs (Scrum Inc.) for providing guidance on how Scrum can be incorporated into the Home Hospital Early Adopters Accelerator. The authors acknowledge Margaret Ben-Or (Ariadne Labs) for early program launch efforts and project managers Adam Linsley (Ariadne Labs) and Carol Lucey (Ariadne Labs). The authors acknowledge Dr. Carme Hernandez (Clinic Barcelona and Brigham and Women’s Hospital) and Dr. Bruce Leff (John Hopkins University) for providing subject matter expertise to participants intermittently throughout the Accelerator.

## Additional File

Additional file 1.docx

- Additional file 1

