## Appendix for "Implementation of Agile in health care: Methodology for a multi-site home hospital accelerator"

### eAppendix

### eAppendix 1

#### Scrum glossary

| Term | Explanation |
|---|---|
| Scrum | Scrum is a lightweight framework that helps people, teams and organizations generate value through adaptive solutions for complex problems. |
| Agile | An organization's ability to adapt based on market changes for their competitive advantage |
| User story | A user story is the smallest unit of work expressed from the user's perspective. The purpose of a user story is to outline in simple language the desired outcome |
| Sprint | Scrum Event that is time-boxed to one month or less, that serves as a container for the other Scrum events and activities. Sprints are done consecutively, without intermediate gaps |
| Scrum Events | Scrum combines four formal events for inspection and adaptation within a containing event, the Sprint. These events work because they implement the empirical Scrum pillars of transparency, inspection, and adaptation. |
| Scrum Artifacts | Scrum's artifacts represent work or value. They are designed to maximize transparency of key information. Thus, everyone inspecting them has the same basis for adaptation.<br>Each artifact contains a commitment to ensure it provides information that enhances transparency and focus against which progress can be measured:<br>• For the Product Backlog it is the Product Goal.<br>• For the Sprint Backlog it is the Sprint Goal.<br>• For the Increment it is the Definition of Done.<br>These commitments exist to reinforce empiricism and the Scrum values for the Scrum Team and their stakeholders. |
| Products Backlog | A Scrum Artifact that consists of an ordered list of the work to be done in order to create, maintain and sustain a product. Managed by the Product Owner. |

| | |
|---|---|
| Sprint Backlog | Scrum Artifact that provides an overview of the development work to realize a Sprint's goal, typically a forecast of functionality and the work needed to deliver that functionality. Managed by the Developers. |
| Product increment | Scrum Artifact that defines the complete and valuable work produced by the Developers during a Sprint. The sum of all Increments form a product. |
| Sprint planning | Scrum Event that is time-boxed to 8 hours, or less, to start a Sprint. It serves for the Scrum Team to inspect the work from the Product Backlog that's most valuable to be done next and design that work into Sprint backlog. |
| Daily Scrum | Scrum Event that is a 15-minute time-boxed event held each day for the Developers. The Daily Scrum is held every day of the Sprint. At it, the Developers plans work for the next 24 hours. This optimizes team collaboration and performance by inspecting the work since the last Daily Scrum and forecasting upcoming Sprint work. The Daily Scrum is held at the same time and place each day to reduce complexity. |
| Sprint Review | Scrum Event that is set to a time-boxed of 4 hours, or less, to conclude the development work of a Sprint. It serves for the Scrum Team and the stakeholders to inspect the Increment of product resulting from the Sprint, assess the impact of the work performed on overall progress toward the Product Goal and update the Product backlog in order to maximize the value of the next period. |
| Sprint Retrospective | Scrum Event that is set to a time-box of 3 hours, or less, to end a Sprint. It serves for the Scrum Team to inspect the past Sprint and plan for improvements to be enacted during future Sprints. |
| Definition of Done | A formal description of the state of the Increment when it meets the quality measures required for the product. The moment a Product Backlog item meets the Definition of Done, an Increment is born. The Definition of Done creates transparency by providing everyone a shared understanding of what work was completed as part of the Increment. If a Product Backlog item does not meet the Definition of Done, it cannot be released or even presented at the Sprint Review. |

#### eAppendix 2

**Developer experience feedback survey questions**

1.The scrum team worked well together

| | | | | |
|---|---|---|---|---|
| Strongly disagree | Disagree | Neutral | Agree | Strongly agree |

2.This scrum sprint was productive

| | | | | |
|---|---|---|---|---|
| Strongly disagree | Disagree | Neutral | Agree | Strongly agree |

3.The scrum team spent an acceptable amount of time working on the product

| | | | | |
|---|---|---|---|---|
| Strongly disagree | Disagree | Neutral | Agree | Strongly agree |

4.This sprint produced a high-quality product

| | | | | |
|---|---|---|---|---|
| Strongly disagree | Disagree | Neutral | Agree | Strongly agree |

#### eAppendix 3

Interview guide

1. What was your experience like participating in the early adopter's accelerator program?
    a. What went well?
    b. What could be improved?

2. What did you think about the 2-week sprints?

   a. Did the process feel rushed, or did it feel like enough time to develop a product?

3. What have you learned from participating in the accelerator?

4. What were your expectations for the products created during the accelerator?
   a. Did the products meet your expectations?

5. How would you define a high-quality product in this context?
   a. Do you think that your scrum team produced a high-quality product(s)? Why or why not?

6. Do you think the product(s) that you created during the program will be feasible to implement at your site? Why or why not?
   a. Do you feel that you have enough time to implement the product (s)
   b. Do you feel that you have the right infrastructure to implement the product (technical infrastructure, operational, enough staff to implement the product etc.)

7. Would you recommend the accelerator program to your colleagues or to another hospital/ health system? Why or why not?

8. Are there any topics that were not covered in this program that you would be interested in working on in the future?

*For sites that have an established home hospital program*

9. In addition to a scrum sprint, what kind of support or programs would have helped your home hospital program start up faster?
   a. What policies (if any) have been put into place at your site that supported the start up of your home hospital program?

*For sites that have not started admitting patients to their home hospital program:*

10. What kind of support or programs would help your home hospital start up faster?
